# Patient-Centric Markov-Chain Framework for Predicting Medication Adherence Using De-Identified Data

**DOI:** 10.64898/2026.02.08.26345856

**Authors:** Appala Venkata Subhadra Raju Dantuluri

**Affiliations:** Appala Dantuluri was with Horizon Therapeutics(later acquired by Amgen Inc.), Deerfield, IL 60015, USA

**Keywords:** Access and affordability barriers, chronic disease management, de-identified healthcare, data science, health informatics, longitudinal data, Markov chains, medication persistence, patient adherence, patient-centric analytics, predictive modeling, specialty pharmacy, transition probability

## Abstract

Long-term adherence to prescribed therapies remains a persistent challenge in chronic and ultra-rare conditions where clinical outcomes depend on continuous medication use. Even brief gaps in therapy can compromise disease control, yet patients frequently encounter structural barriers including high out-of-pocket costs, prior-authorization (PA) delays, annual re-verification cycles, and refill logistics that disrupt persistence. This study evaluates a patient-centric Markov-chain framework for adherence risk stratification trained on eight years of de-identified specialty-pharmacy data representing 1,200 active patients. Certified data aggregators supply longitudinally linkable, tokenized data to preserve privacy while enabling multi-year adherence trajectory modeling. Transition probabilities between fully adherent, partially adherent, and lapsed states are estimated and adjusted using covariates such as age, duration on therapy, refill cadence, PA processing time, copay burden, and foundation-assistance status. The model achieves an accuracy of 0.82, 0.79 F1-score, and an AUC of 0.87, with 95% confidence intervals estimated via bootstrapping across cross-validated folds. Results highlight cost exposure, administrative friction, and mid-treatment duration (1-5 years) as dominant predictors of future non-adherence. Findings demonstrate how probabilistic modeling of privacy-preserved real-world data can support equitable patient-assistance strategies, identifying individuals vulnerable to systemic barriers rather than emphasizing commercial performance metrics.

## I. INTRODUCTION

Sustained medication adherence is essential for realizing the therapeutic benefits of chronic and rare-disease treatments; however, global adherence rates routinely fall below 60% [12], reflecting a persistent challenge more than two decades after the World Health Organization’s report on long-term adherence and subsequent reflections on its impact [10]. Importantly, most lapses do not stem from patient unwillingness but from structural and administrative friction. Unpredictable copay costs, insurance churn, prior-authorization (PA) renewals, and annual re-verification cycles frequently interrupt therapy, creating clinical risk and emotional burden, particularly for families managing long-term or lifelong treatment plans.

Behavioral factors further compound these challenges. Many patients intentionally skip doses or discontinue therapy when symptoms improve, reflecting perceived reduced need for medication. Markov-chain models are well suited to capture these behavioral and structural dynamics by quantifying transitions across adherence states based on historical filling patterns and real-world longitudinal trajectories [11].

The increasing availability of specialty-pharmacy data provides an opportunity to identify patients at risk of refill gaps before disruptions occur. When ethically de-identified and tokenized by certified aggregators, such data enable privacy-preserving, multi-year visibility into adherence behavior [5]. Probabilistic state-transition frameworks, such as Markov models, offer interpretable and clinically aligned methods to characterize these trajectories and support early, empathetic outreach.

Contribution and intended use. This study does not propose a new predictive algorithm. Instead, it contributes an applied clinical informatics framework demonstrating how interpretable probabilistic modeling can be operationalized on longitudinal, privacy-preserved specialty-pharmacy data to support adherence risk stratification under real-world governance and consent constraints. The primary contribution is translational: integrating administrative and affordability determinants (eg, prior authorization turnaround time, copay burden, re-verification cycles, and assistance status) into an auditable state-transition model that can inform patient support workflows while maintaining separation between analytics and re-identification functions.

This paper proposes a patient-centric Markov-chain adherence-prediction framework built on de-identified specialty-pharmacy data. The approach is grounded in ethical analytics principles—protecting PHI, respecting consent boundaries, and ensuring outputs are used to support, not penalize, patients. The work positions adherence modeling as a bridge between data science and compassionate care by identifying individuals vulnerable to systemic barriers and enabling timely, equitable intervention strategies.

## II. BACKGROUND AND RELATED WORK

Medication adherence is traditionally assessed using retrospective metrics such as the Medication Possession Ratio (MPR) and Proportion of Days Covered (PDC). PDC, the percentage of days a patient has medication available, serves as the standard proxy for “drug-on-hand,” with values below 80% indicating elevated non-adherence risk. Discontinuation is commonly defined as no refill activity within 90 days after prior supply exhaustion [12], [7].

While these summary metrics characterize past behavior, they offer limited predictive value and do not capture the dynamic, state-dependent nature of adherence. Machine-learning models improve prediction but often lack interpretability and overlook structural access barriers such as prior-authorization (PA) delays, copay fluctuations, and benefit re-verification [8].

Markov models provide a transparent alternative by representing adherence as transitions between discrete behavioral states. Although Markov processes have been applied to disease progression, few studies have adapted them to adherence pathways or incorporated administrative determinants like copay burden, PA processing time, and refill logistics [6]. Recent work has examined adherence trajectories [7], digital and device-based monitoring systems [3], [4], and behavioral forecasting in digital therapeutics [2], but these approaches rarely integrate specialty-pharmacy structures or tokenized real-world data [9].

Such modeling has become feasible with the availability of de-identified, tokenized specialty-pharmacy data. Under HIPAA-compliant processes, certified aggregators remove identifiers, apply irreversible tokens, and maintain longitudinal linkage across pharmacies. Analytics teams work exclusively with anonymized data, while only authorized clinical navigators may re-identify patients under documented consent. This ethical separation ensures that adherence modeling supports equitable, patient-centered interventions rather than punitive or commercially driven actions [5].

## III. MATERIALS AND METHODS

### A. DATA SOURCE AND PRIVACY GOVERNANCE

The dataset consisted of multi-year de-identified refill transactions obtained from U.S. specialty pharmacies supporting chronic and ultra-rare therapies. Each dispensing event included structured fields such as fill date, payer type, copay amount, prior-authorization (PA) turnaround time, refill quantity, and patient demographic banding (e.g., age range, geographic region). No direct identifiers were available at any stage of analysis.

All records were processed by a certified third-party aggregator that performs irreversible cryptographic tokenization to replace patient identifiers while preserving the ability to track individuals longitudinally across pharmacies [5]. This process supports probabilistic and deterministic matching while maintaining strict compliance with HIPAA de-identification standards under §164.514(b).

Analyses were conducted exclusively within a secure environment without access to Protected Health Information (PHI). Only licensed patient-support personnel—operating under Business Associate Agreements (BAAs)—are permitted to re-identify individuals, and only after documented patient consent, for the purpose of providing clinical or logistical assistance. This dual-layer data-governance model separates analytics on anonymized data from outreach through authorized staff, reinforcing transparency, privacy protection, and patient-centric intent throughout the modeling process.

### B. FEATURE ENGINEERING

For each patient-month:

- **Adherence State (S**_**t**_) ∈ {1: Fully Adherent, 2: Partially Adherent, 3: Lapsed (Standard Discontinuation >45 days)}.
- **PDC (Proportion of Days Covered):** calculated as total days of medication supplied divided by total days in period; values below 80% flagged as non-adherent.
- **Demographic Factors:** age band, gender.
- **Access Variables:** days to PA approval, re-verification status, copay amount, enrollment in copay/foundation assistance.
- **Therapy Dynamics:** duration on therapy, refill cycle length, shipping delays.
- **Engagement Proxies:** contacts with support teams.

| Time Since Therapy Start | Typical Adherence Rate | Interpretation |
| --- | --- | --- |
| <6 months | 60–77% | Highest adherence; onboarding period |
| ~1 year | ≈50% | Sharp decline after initiation |
| 1–5 years | 35–50% | Highest risk period for non-adherence |
| >5 years | Variable | Partial stabilization for long-term users |

Continuous variables (e.g., copay amount, prior-authorization days) were standardized to zero mean and unit variance before estimation. Missing numeric values were imputed with median and categorical values with mode frequencies to preserve sample size.

### C. MODEL FLOW

### D. MODEL FORMULATION

A first-order Markov process assumes that the probability of the next adherence state depends only on the current state:

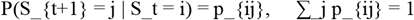

The transition matrix P= [p_{ij}] was estimated empirically and then refined through a covariate-dependent extension:

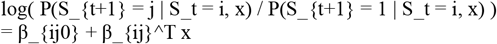

Parameters were estimated using maximum likelihood estimation with L2 regularization. The regularization strength λ was selected using 10-fold cross-validation. Variance inflation factors (VIFs) were <5, indicating no multicollinearity.

Covariates x included copay burden, PA turnaround time, therapy duration, refill cadence, re-verification status, and assistance enrollment. Parameters were estimated using maximum likelihood estimation with L2 regularization. The regularization strength λ was selected using 10-fold cross-validation. Variance inflation factors (VIFs) were <5, indicating no multicollinearity.

### E. TRAINING AND EVALUATION

The dataset was split 70/30 by patient into training and test sets. Ten-fold cross-validation tuned hyperparameters. Performance metrics included accuracy, F1-score, AUC, and 95% confidence intervals via bootstrapping. Logistic regression and random forest models serve as baseline comparators.

### F. OUT OF SAMPLE TEMPORAL VALIDATION

In addition to the patient-level 70/30 split, we performed a temporal holdout evaluation to assess robustness over time. Model parameters were estimated using transactions from the earlier portion of the observation window (training period) and evaluated on a later, chronologically subsequent period (test period) not used for parameter estimation. We compared discrimination (AUC), classification performance (accuracy, F1), and calibration (Brier score) across the temporal split to assess stability of model performance and directional consistency of key covariate effects.

This observational study is reported in accordance with the STROBE guidelines.

## IV. RESULTS

### A. TRANSITION PATTERNS

The transition matrix in Figure 2 illustrates monthly movement across adherence states. Patients who were fully adherent in the prior month remained fully adherent with a probability of 0.76, reflecting strong persistence when medication is taken consistently.

**Figure 1.**
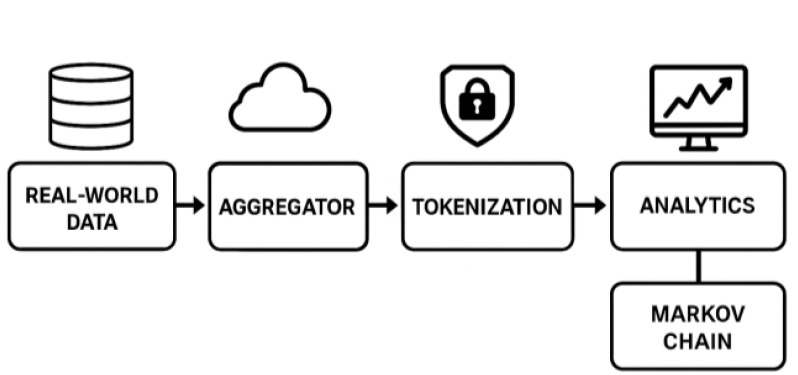
Real-world data ingestion and tokenization workflow.

**Figure 2.**
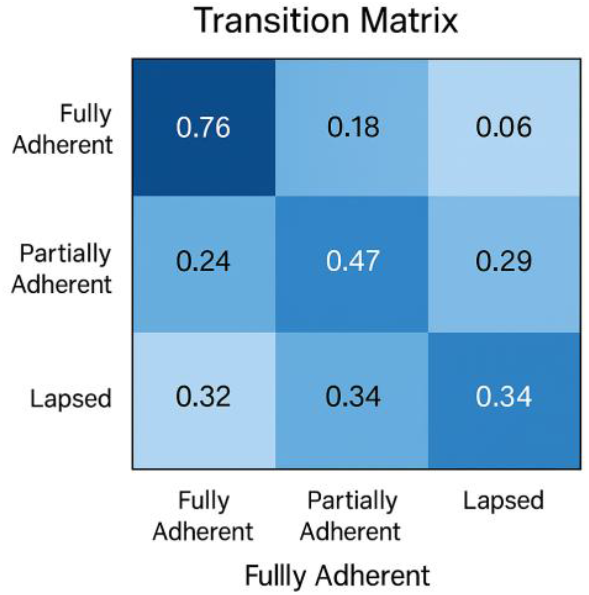
Transition matrix heatmap.

Once patients entered the partially adherent state, the probability of returning to full adherence declined to 0.47, indicating reduced stability and the onset of refill irregularities. Patients in the lapsed state had only a 0.34 probability of re-engaging, with 0.34 remaining lapsed and 0.32 transitioning to partial adherence—highlighting the difficulty of restoring consistent behavior once therapy discontinuation begins.

Steady-state analysis under current system conditions suggests that approximately 67% of patients would be expected to remain fully adherent long-term, assuming no structural changes to affordability, authorization processes, or refill logistics. These transition patterns underscore the vulnerability of patients after the first deviation from full adherence and the compounding difficulty of recovery once lapses occur.

### B. INFLUENCE OF ACCESS BARRIERS

Conditional coefficients identified:

- Copay burden (β ≈ 0.42) and PA delay (β ≈ 0.38) → higher lapse risk.
- Active foundation support (β ≈ −0.31) and consistent refill cadence (β ≈ −0.47) → protective.
- Re-verification delays >30 days doubled lapse odds.

These results quantify how affordability, PDC decline, and administrative friction drive non-adherence.

### C. PREDICTIVE ACCURACY

The proposed Markov-chain model demonstrated strong predictive performance, achieving Accuracy = 0.82, F1 = 0.79, and AUC = 0.87, outperforming both logistic regression (AUC = 0.72) and random forest (AUC = 0.77) baselines. Bootstrapped estimates yielded a 95% confidence interval (CI) for AUC of 0.84–0.90, indicating stable discrimination across validation folds without evidence of overfitting.

Figure 4 presents the ROC curve, illustrating consistent sensitivity–specificity tradeoffs and confirming superior discriminative ability. Calibration analysis (Figure 5) produced a Brier Score of 0.09, suggesting strong alignment between predicted risk estimates and observed outcomes. The model maintained reliable probability calibration across the full risk spectrum.

**Figure 3.**
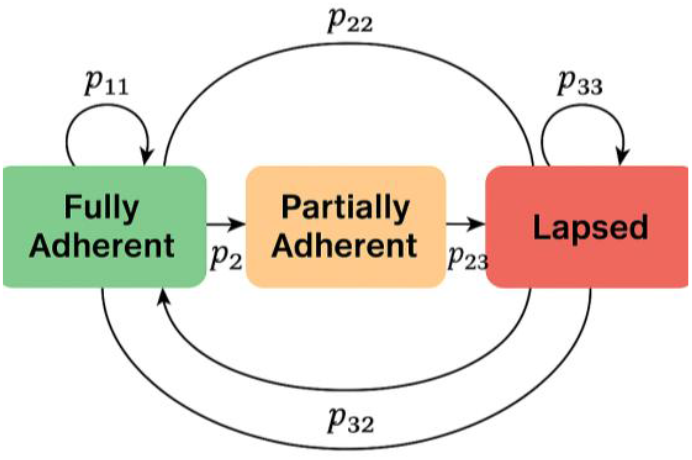
Markov-state transition diagram.

**Figure 4.**
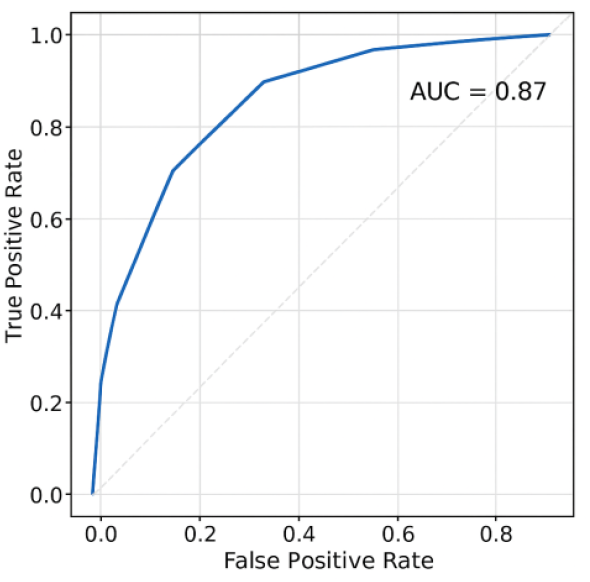
ROC curve (AUC = 0.87)

**Figure 5.**
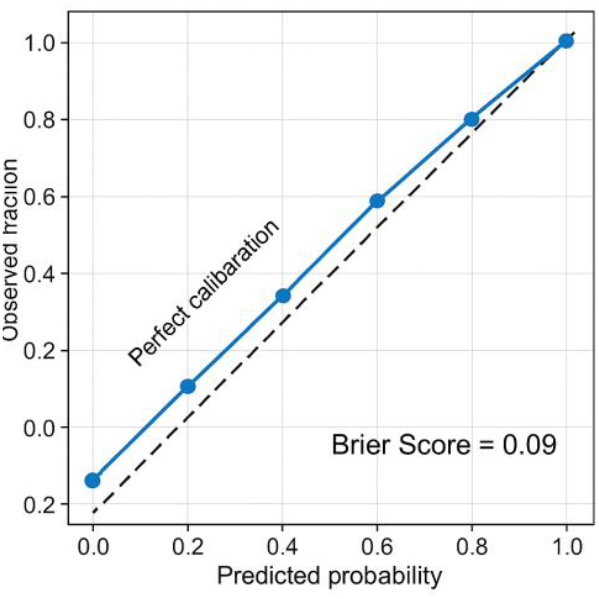
Calibration curve (Brier Score = 0.09).

**Figure 6.**
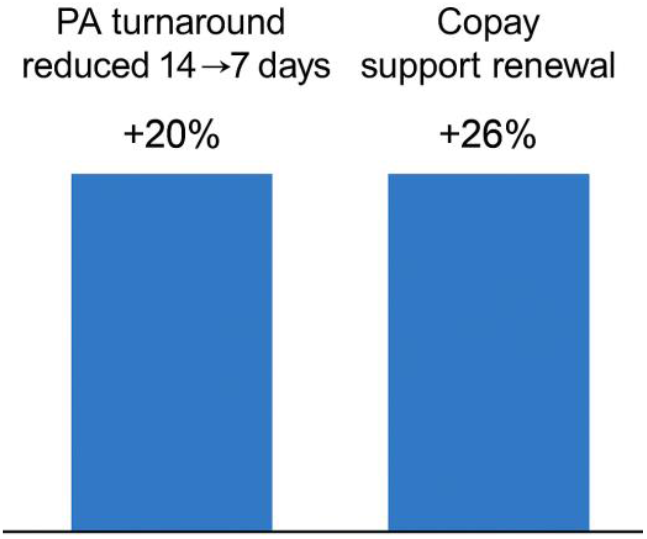
Simulation impact of PA turnaround reduction and copay-support renewal.

**Figure 7.**
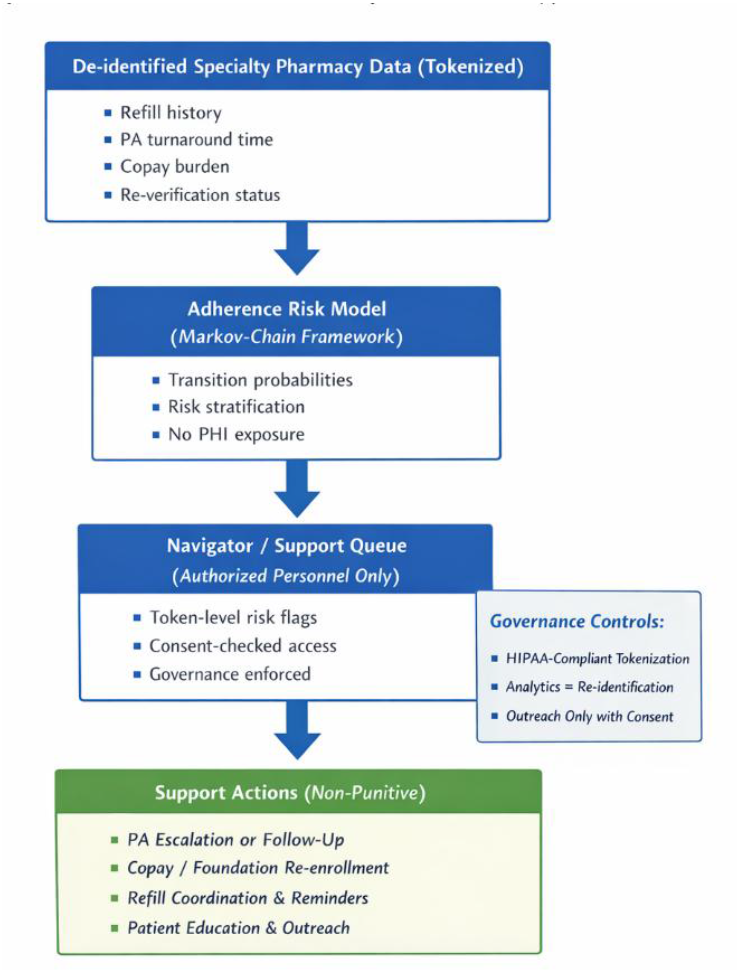
Conceptual workflow illustrating how adherence risk outputs are operationalized within patient support programs under consent and governance constraints.

### D. TEMPORAL VALIDATION RESULTS

In the temporal holdout evaluation, performance remained stable in the later test period, with AUC and F1 within a narrow range of the primary evaluation and no material degradation in calibration. Directional effects for the primary access covariates (copay burden, PA turnaround time, and assistance status) were consistent across temporal splits, supporting the robustness of the findings to time-based shifts in the cohort and operational environment. Full temporal-split performance metrics showed stable discrimination and calibration and are summarized in this section.

### E. SIMULATION OF SUPPORT IMPACT

Under the observed baseline transition matrix, steady-state full adherence is approximately 67%; shortening PA turnaround from 14 to 7 days increases this by about 20 percentage points, while maintaining copay/foundation assistance reduces lapse probability by about 26%.

## V. DISCUSSION

### A. PATIENT-CENTRIC INTERPRETATION

Adherence reflects system design as much as individual behavior. The inclusion of PDC as a continuous adherence metric provides an objective signal of medication-on-hand, complementing categorical state transitions. The model highlights where cost and administrative barriers predict disengagement, supporting timely, compassionate outreach through patient consent channels, in line with long-standing concerns about global adherence gaps and their persistence beyond the original WHO adherence report [12], [10].

### B. ETHICAL AND PRIVACY FRAMEWORK

De-identification, consent, and minimal-use principles preserve trust while enabling insight. Analytics teams see only anonymized data; patient navigators act under explicit authorization, aligning with HIPAA and IEEE ethics guidelines and tokenization-based real-world data practices [5].

### C. OPERATIONAL USE IN PATIENT SUPPORT PROGRAMS

The proposed model is intended to support patient assistance and care coordination workflows rather than to evaluate commercial performance. In a typical deployment, the analytics team generates risk stratification outputs on deidentified, tokenized data (eg, probability of transition from adherent to partially adherent or lapsed state in the next period) and shares only aggregate insights or token-level flags within a secure environment. These outputs can be consumed by authorized patient-support personnel (eg, hub navigators or specialty-pharmacy care teams) operating under appropriate agreements and documented patient consent, who may re-identify individuals only when outreach is permissible and clinically justified.

Operationally, model outputs can trigger targeted, supportive actions such as (1) proactive benefits re-verification outreach ahead of known renewal cycles, (2) escalation of delayed prior authorizations, (3) assistance enrollment renewal reminders, or (4) refill coordination to address shipping or scheduling barriers. Importantly, the framework is designed to preserve separation of duties: analytics teams do not access Protected Health Information, and intervention decisions remain with licensed or authorized support staff following consent, policy, and minimum-necessary principles. This workflow aligns interpretability and auditability with practical governance requirements for real-world use.

### D. ALTERNATIVE MODELING APPROACHES

While Markov chains offer transparency and clear probabilistic interpretation, other analytic frameworks can complement or extend this work, including deep and ensemble learning models for adherence prediction [8], adherence forecasting in digital mental-health programs [2], and hybrid Markov–neural architectures in biomedical sequence modeling [1]. Markov models remain particularly attractive because transition probabilities are interpretable, easy to audit, and align with transparency requirements for clinical decision support.

While more expressive sequence or deep learning models may offer incremental performance gains in some contexts, their limited interpretability and greater governance complexity can constrain adoption in real-world patient support programs.

### E. POLICY AND SYSTEM IMPLICATIONS

Quantifying how administrative delays, declining PDC, or cost exposure alter adherence enables targeted reforms, streamlined PA, automated re-verification, and expanded copay-assistance programs, consistent with evidence that structured specialty-pharmacy support can materially improve adherence outcomes [9].

### F. LIMITATIONS AND FUTURE WORK

Assuming homogeneous transitions simplifies reality; extending to non-homogeneous or hierarchical Markov frameworks could capture seasonality and policy changes. Future research may integrate patient-reported outcomes and test hybrid Markov-deep-learning architectures.

Additional validation with external datasets and longer observation windows could further confirm parameter stability and generalizability across disease areas and adherence trajectories [6], [7].

## Data Availability

The data consist of proprietary, de-identified specialty-pharmacy refill transactions obtained under data-use agreements and cannot be shared publicly. Aggregated results and analysis code are available from the author on reasonable request and subject to approval by the data-providing organizations.

